# Associations between reported healthcare disruption due to COVID-19 and avoidable hospitalisation: Evidence from seven linked longitudinal studies for England

**DOI:** 10.1101/2023.02.01.23285333

**Authors:** Mark A. Green, Martin McKee, Olivia Hamilton, Richard J. Shaw, John Macleod, Andy Boyd, The LH&W NCS Collaborative, and Srinivasa Vittal Katikireddi

## Abstract

**Background:** Health services across the UK struggled to cope during the COVID-19 pandemic. Many treatments were postponed or cancelled, although the impact was mitigated by new models of delivery. While the scale of disruption has been studied, much less is known about if this disruption impacted health outcomes. The aim of our paper is to examine whether there is an association between individuals experiencing disrupted access to healthcare during the pandemic and risk of an avoidable hospitalisation.

**Methods:** We used individual-level data for England from seven longitudinal cohort studies linked to electronic health records from NHS Digital (n = 29 276) within the UK Longitudinal Linkage Collaboration trusted research environment. Avoidable hospitalisations were defined as emergency hospital admissions for ambulatory care sensitive and emergency urgent care sensitive conditions (1^st^ March 2020 to 25^th^ August 2022). Self-reported measures of whether people had experienced disruption during the pandemic to appointments (e.g., visiting their GP or an outpatient department), procedures (e.g., surgery, cancer treatment) or medications were used as our exposures. Logistic regression models examined associations.

**Results:** 35% of people experienced some form of disrupted access to healthcare. Those whose access was disrupted were at increased risk of any (Odds Ratio (OR) = 1.80, 95% Confidence Intervals (CIs) = 1.34-2.41), acute (OR = 1.68, CIs = 1.13-2.53) and chronic (OR = 1.93, CIs = 1.40-2.64) ambulatory care sensitive hospital admissions. There were positive associations between disrupted access to appointments and procedures to measures of avoidable hospitalisations as well.

**Conclusions:** Our study presents novel evidence from linked individual-level data showing that people whose access to healthcare was disrupted were more likely to have an avoidable or potentially preventable hospitalisation. Our findings highlight the need to increase healthcare investment to tackle the short- and long-term implications of the pandemic beyond directly dealing with SARS-CoV-2 infections.

## Introduction

The COVID-19 pandemic created unprecedented disruption to healthcare in the UK. Health facilities reoriented to care for surging numbers of patients with COVID-19, initially through postponing or cancelling non-emergency treatment and diagnostic tests. People were deterred from seeking healthcare by fear of being exposed to SARS-CoV-2 in health facilities (1), altruistic behaviours aimed at ‘protecting’ the NHS, and reduced availability of face-to-face consultations (2). Collectively, these phenomena have resulted in fewer GP consultations (3), diagnostic tests (4), cancer referrals, diagnoses and treatments (3,5,6), elective and emergency hospital admissions (7,8), and increased waiting times to initiate treatment (9). Although these impacts are not unique to the UK (10,11), it has fared much worse than many otherwise similar countries and is unique among industrialised countries in failing to bring people back into the workforce post-pandemic (12), potentially because of persisting high levels of ill health and unmet need for care (13).

The extent of healthcare disruption has been described elsewhere (14), but to our knowledge this has not been linked to empirically observed adverse health outcomes at the individual level, even though the risks are clear, with delays in diagnosis and treatment allowing illnesses to progress to greater severity. One study has modelled the potential consequences of delays to cancer diagnoses, applying historic data to pandemic scenarios. The authors estimated that the resulting disruption will result in between 3,291 and 3,621 avoidable deaths in England from the four most common cancers (Breast, Colorectal, Lung and Oesophageal) within 5 years (15). We urgently need empirical evidence on the scale and nature of the impact of the pandemic on healthcare disruption to identify if we need to respond to its consequences, and where any responses should be focused.

Understanding the impacts of disrupted access to health is, however, difficult due to the many elements of different care pathways, each with different impacts on health outcomes. To make the effects of this disruption observable, we employ a concept from the health systems literature. Avoidable hospitalisations are unplanned admissions that could potentially have been prevented through timely care delivered in the community. The concept is used as a ‘warning sign’ for failings in health system performance and is a key metric used in the NHS (16–19). We hypothesize that people whose care was disrupted during the pandemic would be more likely to have an avoidable hospitalisation. Given that pandemic-disruption has affected the lives of everyone, this approach allows us to evaluate the overall impact of disruption rather than focusing on discrete services whose study may obscure the overall effect of society-wide disruption.

It is very difficult to capture the individual experience of healthcare disruption from electronic health records, but it can be identified in longitudinal surveys. By linking data on participants in longitudinal cohorts with their electronic health records, we can describe the impact of any disruption they experience on health outcomes at an individual level. We are not aware of any research that has used linked individual-level data to study the effects of COVID-19 healthcare disruption.

The aim of our paper is to examine whether there is an association between individuals experiencing disrupted access to healthcare during the pandemic and risk of an avoidable hospitalisation.

## Methods

### Data

We used data on individuals from seven UK population-based longitudinal studies linked to electronic health records from NHS Digital for England. These include five birth cohorts (1946 National Survey of Health and Development, 1958 National Child Development Study, 1970 British Cohort Study, Next Steps and Millennium Cohort Study) and two age-heterogenous studies (English Longitudinal Study of Ageing and Understanding Society). Each cohort is described in Appendix Table K. The cohort data were accessed using the UK Longitudinal Linkage Collaboration (UK LLC). The UK LLC trusted researcher environment hosts de-identified data from many longitudinal population studies and systematically links these to participants’ health, administrative and environmental records within a secure analysis environment. Ethical approval for the project was granted by the University of Liverpool’s Research Ethics Board [reference 10634].

Each cohort study sent surveys to members of their cohorts inquiring about their experiences during the COVID-19 pandemic, supplementing their existing data collection processes. We use these data here. The data from all cohorts were pooled, giving a total sample size of 41 439. Combining cohorts brings value through improving the representativeness of the data and increasing statistical power (14). We excluded people residing outside England as data linkage was not possible (n = 5975). We further excluded individuals who did not consent to linkage or for whom linkage was not possible (n = 5911). Participants with linked data experienced more disruption than those with unlinked data, although differences were only small (Appendix Table A). We excluded all individuals who died during the study period (n = 277). The total analytical sample size was n = 29 276 (Appendix Table B breaks down sample sizes by cohort).

### Outcomes

Linkage of cohort members to electronic health records was conducted by the UK LLC. Electronic health records from NHS Digital included civil registration of deaths, secondary care (hospital episode statistics admitted patient care), and vaccination status. We selected records between 1^st^ March 2020 (which we define as the start of NHS disruption) and 25^th^ August 2022 (end of available data), which thus comprise the study period.

We selected two measures of unplanned avoidable hospitalisations commonly used for evaluating NHS performance: Ambulatory Care Sensitive (ACSC) and Emergency Urgent Care Sensitive (EUCS) conditions (16,17). ACS are conditions that can be, in theory, treated through community care and therefore should not require hospital admission (17,20). We use an overall measure for any ACSC, as well as stratify by ACSC type into (i) acute (e.g., cellulitis, dental caries, rickets, gastric ulcer), (ii) chronic (e.g., hypertension, angina, asthma), (iii) vaccine-preventable conditions (e.g., mumps, measles, influenza). EUCS are acute exacerbations of urgent conditions that will potentially result in hospital admission, but that the NHS should be trying to treat within the community to minimise the need for hospital care (17,20). Code lists were designed to match NHS Digital’s approach (openly available at https://www.opencodelists.org/users/mgreen/) and we selected any emergency hospital admission during the study period where codes were present in the primary diagnosis field (position 1). We also derive an outcome of whether an individual had any hospital admission during the study period to contextualise our findings. All outcome variables in the main analysis were binary outcomes.

We also used COVID-19 vaccination status (binary: individual had received two COVID-19 doses by the end of the study period or not) as a falsification test (21,22). We did not expect there to be an association between experiences of healthcare disruption and vaccination uptake, because vaccine delivery was prioritised and therefore less disrupted. It therefore provides an imperfect, but valuable, instrumental indicator to assess the role of unobserved confounding in our models.

### Exposures

Our primary exposure variable was whether individuals self-reported any disruption to healthcare (i.e., cancelled or postponed care, changes to planned/existing treatments). This was measured across all waves of data collection. We further stratified our exposure by type of disruption into (i) appointments (e.g., visiting their GP or an outpatient department), (ii) procedures (e.g., surgery, cancer treatment), and (iii) medications. This allows us to examine the different pathways through which disruption affected individuals. Descriptions of questions asked in surveys are reported in Appendix Table C.

### Control variables

We avoided over-adjustment by controlling only for key confounding variables. Variables were selected from those consistent across cohorts, limiting the measures we could include. Personal characteristics of age (numeric), sex (male or female), and ethnicity (White or racially minoritized group) were included to account for demographic differences. The inconsistent ethnic categorisations used meant that we had to amalgamate into a simplistic binary definition. Self-rated health status (excellent/good or fair/poor) was included since we hypothesised that individuals with poor health were more likely to experience disruption and hospitalisations. While self-rated health status may sit on our causal pathway between disruption and health outcomes (e.g., people with poor health were more vulnerable to the effects of disruption and risk of an avoidable hospitalisation), excluding it from our models did not lead to materially different findings. Socioeconomic position was measured as housing tenure (owned house outright/with mortgage or not) and neighbourhood socioeconomic deprivation (2019 index of multiple deprivation quintile provided via linkage by NHS Digital). We also adjusted for the longitudinal cohort that individuals were in (categorical variable). We selected the most recent value for each measure during the COVID-19 waves.

### Statistical analyses

Descriptive statistics were calculated to provide summary measures of our data. Logistic regression models were used with unadjusted (exposures only) and adjusted (exposures and control variables) models presented here. In the main analysis, we considered any outcome during the study period since we are unsure when experiences of disruption occurred (individuals were only asked to report if they had experienced disruption at any point). Two sensitivity analyses were undertaken to assess how robust this model specification was. First, we considered outcomes which took place after the final survey date so that we are certain that any disruption occurred before outcomes (same logistic regression model used). Second, we used a Cox regression model for time to outcome from the last survey date.

Although this model specification is not time since the exposure, it may be better at handling rarer outcomes. All analyses were adjusted for the sampling design of each survey, including sample weights that account for representativeness, attrition, and non-response (i.e., sample weights, primary sampling units, strata and finite population correction factor were adjusted for). The numbers of missing data across our variables are presented in Appendix Table D. Missing values for each variable were imputed using polytomous regression using all other exposure and control variables. All analyses were conducted using R statistical software and the code is openly available (https://github.com/UKLLC/LLC_0009).

## Results

Table 1 presents summary statistics of our analytical sample. Each of our outcomes was uncommon during our study period. By 25^th^ August 2022, 14% (weighted percentage) of participants had a hospital admission. 3% of participants were admitted for an ambulatory care sensitive condition. Among these admissions, vaccine-preventable admissions were the least common (0.8%). 35% of participants reported experiencing any form of disruption in their access to healthcare due to COVID-19. Disruption was most commonly experienced in accessing appointments (26%), followed by procedures (18%). Few individuals experienced disruption in their access to medications (6%). Summary statistics for control variables and sensitivity analyses are presented in Appendix Tables E and F respectively.

**Table 1:** Descriptive statistics for outcome variables and exposures in the pooled sample.

| Measure | Frequency | Unweighted Percentage | Weighted Percentage |
| --- | --- | --- | --- |
| Total admissions | 3618 | 12.36 | 13.65 |
| Ambulatory care sensitive any | 780 | 2.66 | 3.36 |
| Ambulatory care sensitive acute | 347 | 1.19 | 1.32 |
| Ambulatory care sensitive chronic | 369 | 1.26 | 1.39 |
| Ambulatory care sensitive vaccine preventable | 94 | 0.32 | 0.78 |
| Emergency urgent care sensitive | 625 | 2.13 | 2.37 |
| Two COVID-19 vaccine doses | 27513 | 93.98 | 92.64 |
| Disruption any | 9742 | 33.28 | 34.79 |
| Disruption appointments | 7456 | 25.47 | 26.20 |
| Disruption medications | 1568 | 5.36 | 5.86 |
| Disruption procedures | 5292 | 18.08 | 18.12 |

Table 2 presents results from a series of logistic regression models relating experiences of healthcare disruption to measures of avoidable hospitalisations. We found positive associations between healthcare disruption and each outcome in all unadjusted models, although estimates for vaccine-preventable ambulatory care sensitive conditions were imprecise due to few observed events. Following adjustment for known explanatory factors, positive associations where confidence intervals (CIs) did not cross 1 remained for any, acute and chronic ambulatory care sensitive conditions. People who experienced any form of healthcare disruption had 80% higher odds of being admitted to hospital for any ambulatory care sensitive condition (Odds Ratio (OR) = 1.80, 95% CIs = 1.34-2.41), 68% higher odds of being admitted for an acute ambulatory care sensitive condition (OR = 1.68, CIs = 1.13-2.53), and 93% higher odds of being admitted for a chronic ambulatory care sensitive condition (OR = 1.93, CIs = 1.40-2.64). For any hospital admission, we find a positive association in both unadjusted and adjusted analyses. In the adjusted model, individuals who experienced disruption to healthcare had 92% higher odds (OR = 1.92, CIs = 1.65-2.23) of being hospitalised during the study period.

**Table 2:** Model summary statistics for a logistic (binomial) regression exploring associations between experiences of healthcare disruption and whether an individual had an avoidable hospitalisation.

| Model | Odds Ratio | Lower 95% CI | Higher 95% CI | P value |
| --- | --- | --- | --- | --- |
| <u>Any ambulatory care sensitive</u> |  |  |  |  |
| Unadjusted | 2.75 | 1.75 | 4.31 | <0.001 |
| Adjusted | 1.80 | 1.34 | 2.41 | <0.001 |
| <u>Acute ambulatory care sensitive</u> |  |  |  |  |
| Unadjusted | 1.92 | 1.31 | 2.80 | 0.001 |
| Adjusted | 1.68 | 1.13 | 2.53 | 0.011 |
| <u>Chronic ambulatory care sensitive</u> |  |  |  |  |
| Unadjusted | 3.06 | 2.23 | 4.22 | <0.001 |
| Adjusted | 1.93 | 1.40 | 2.64 | <0.001 |
| <u>Vaccine-preventable ambulatory care sensitive</u> |  |  |  |  |
| Unadjusted | 3.97 | 0.85 | 18.54 | 0.079 |
| Adjusted | 1.40 | 0.70 | 2.80 | 0.344 |
| <u>Emergency urgent care sensitive</u> |  |  |  |  |
| Unadjusted | 1.55 | 1.20 | 2.03 | 0.001 |
| Adjusted | 1.07 | 0.81 | 1.42 | 0.625 |
| <u>Any hospital admission</u> |  |  |  |  |
| Unadjusted | 2.64 | 2.32 | 3.03 | <0.001 |
| Adjusted | 1.92 | 1.65 | 2.23 | <0.001 |

Note: Estimates are Odds Ratios. CI = Confidence Interval. Model adjustment includes the following variables: age, sex, ethnicity, housing tenure, self-rated health status, and longitudinal cohort).

We performed two sensitivity analyses. First, we re-ran the same models but excluded any hospital admissions that had occurred before the final survey wave (Appendix Table G). Results were broadly similar to those presented in Table 2, except that confidence intervals with acute ambulatory care sensitive conditions now passed 1. Second, we ran a time-to-event model for outcomes after the last known survey date (Appendix Table H). Results were similar, with positive associations (unadjusted and adjusted) with all outcomes (only vaccine-preventable ambulatory care sensitive conditions had confidence intervals that passed 1).

When stratifying analyses by type of healthcare disruption experienced (Table 3), we obtained inconsistent results (i.e., 59% of associations saw confidence intervals crossing 1). Individuals who experienced disruption to their procedures had increased odds of being hospitalised for chronic ambulatory care sensitive conditions (OR = 2.20, CIs = 1.42-3.46), and for an emergency urgent sensitive condition (OR = 1.54, CIs = 1.07-2.18). There was a negative association to vaccine-preventable ambulatory care sensitive conditions (OR = 0.39, CIs = 0.17-0.90). We also found positive associations between disruption to procedures and appointments and any hospital admission. Individuals who experienced disruption in accessing appointments had 51% higher odds (OR = 1.51, CIs = 1.05-2.14) of a hospital admission for any ambulatory care sensitive condition. Few other associations with this exposure were identified and, where they were found, they were attenuated following adjustment. We found a large, yet imprecisely estimated, positive association between disruption to access of medications and any ambulatory care sensitive condition (OR = 2.72, CIs = 1.20-6.11). We did not find any clear associations between this type of disruption and any other outcomes.

**Table 3:** Model summary statistics for a logistic (binomial) regression exploring associations between experiences of three types of healthcare disruption (procedures, medications and appointments) and whether an individual had an avoidable hospitalisation (by type).

| Model | Odds Ratio | Lower 95% CI | Higher 95% CI | P value |
| --- | --- | --- | --- | --- |
| <u>Any ambulatory care sensitive - unadjusted model</u> |  |  |  |  |
| Appointments | 2.12 | 1.12 | 4.01 | 0.021 |
| Medications | 2.92 | 0.85 | 10.07 | 0.089 |
| Procedures | 1.31 | 0.64 | 2.66 | 0.451 |
| <u>Any ambulatory care sensitive - adjusted model</u> |  |  |  |  |
| Appointments | 1.51 | 1.05 | 2.14 | 0.025 |
| Medications | 2.72 | 1.20 | 6.11 | 0.017 |
| Procedures | 1.27 | 0.85 | 1.90 | 0.236 |
| <u>Acute ambulatory care sensitive - unadjusted model</u> |  |  |  |  |
| Appointments | 1.57 | 1.02 | 2.41 | 0.039 |
| Medications | 1.06 | 0.38 | 2.94 | 0.912 |
| Procedures | 1.48 | 0.99 | 2.20 | 0.055 |
| <u>Acute ambulatory care sensitive - adjusted model</u> |  |  |  |  |
| Appointments | 1.40 | 0.91 | 2.14 | 0.123 |
| Medications | 1.04 | 0.34 | 3.19 | 0.951 |
| Procedures | 1.45 | 0.97 | 2.16 | 0.070 |
| <u>Chronic ambulatory care sensitive - unadjusted model</u> |  |  |  |  |
| Appointments | 1.52 | 0.94 | 2.46 | 0.086 |
| Medications | 0.71 | 0.41 | 1.23 | 0.227 |
| Procedures | 2.94 | 1.80 | 4.85 | <0.001 |
| <u>Chronic ambulatory care sensitive - adjusted model</u> |  |  |  |  |
| Appointments | 1.26 | 0.83 | 1.93 | 0.286 |
| Medications | 0.76 | 0.38 | 1.54 | 0.441 |
| Procedures | 2.20 | 1.42 | 3.46 | <0.001 |
| <u>Vaccine-preventable ambulatory care sensitive - unadjusted model</u> |  |  |  |  |
| Appointments | 4.71 | 1.09 | 20.49 | 0.038 |
| Medications |  |  |  |  |
| Procedures | 0.23 | 0.04 | 1.31 | 0.096 |
| <u>Vaccine-preventable ambulatory care sensitive - adjusted model</u> |  |  |  |  |
| Appointments | 1.51 | 0.83 | 2.77 | 0.177 |
| Medications |  |  |  |  |
| Procedures | 0.39 | 0.17 | 0.90 | 0.026 |
| <u>Emergency urgent care sensitive - unadjusted model</u> |  |  |  |  |
| Appointments | 1.04 | 0.73 | 1.49 | 0.835 |
| Medications | 0.92 | 0.59 | 1.42 | 0.713 |
| Procedures | 1.93 | 1.32 | 2.83 | 0.001 |
| <u>Emergency urgent care sensitive - adjusted model</u> |  |  |  |  |
| Appointments | 0.87 | 0.61 | 1.22 | 0.411 |
| Medications | 0.90 | 0.52 | 1.57 | 0.726 |
| Procedures | 1.54 | 1.07 | 2.18 | 0.019 |
| <u>Any hospital admission - unadjusted model</u> |  |  |  |  |
| Appointments | 1.49 | 1.23 | 1.80 | <0.001 |
| Medications | 0.90 | 0.64 | 1.26 | 0.524 |
| Procedures | 2.27 | 1.86 | 2.77 | <0.001 |
| <u>Any hospital admission - adjusted model</u> |  |  |  |  |
| Appointments | 1.26 | 1.02 | 1.54 | 0.029 |
| Medications | 0.97 | 0.58 | 1.62 | 0.911 |
| Procedures | 1.93 | 1.55 | 2.41 | <0.001 |

Note: Estimates are Odds Ratios. CI = Confidence Interval. Model adjustment includes the following variables: age, sex, ethnicity, housing tenure, whether had COVID-19, self-rated health status, and longitudinal cohort). Results for vaccine-preventable ambulatory care sensitive conditions and disruption to medications were not robust due to small number issues.

In our sensitivity analyses, we excluded any hospital admissions that had occurred before the final survey wave and then ran logistic regression (Appendix Table I) and Cox regression (Appendix Table J) models. While most associations were consistent in the logistic regression model, there were some differences. There was an additional positive association between disruption to procedures and acute ambulatory care sensitive conditions. Disruption to appointments were all non-significant in adjusted models. In the Cox regression model we found a greater number of positive associations between our variables where confidence intervals were both above 1, including for vaccine-preventable ambulatory care sensitive conditions and disruption to procedures.

Finally, we investigated whether our measures of healthcare disruption were associated with COVID-19 vaccination uptake as a falsification test (Table 4). Looking at overall experiences of healthcare disruption, we found a positive association in unadjusted analyses which was attenuated post-adjustment. When considering type of healthcare disruption, associations for disruption to medications and procedures saw small magnitude and confidence intervals crossed 1. We found a positive association for disruption to appointments that was attenuated following adjustment. This suggests low risk of bias in our fully adjusted associations.

**Table 4:** Model summary statistics for a logistic (binomial) regression exploring associations between experiences of healthcare disruption (separate models for different exposure specifications) and whether an individual was fully vaccinated or not.

| Model | Odds Ratio | Lower 95% CI | Higher 95% CI | P value |
| --- | --- | --- | --- | --- |
| <u>Any healthcare disruption</u> |  |  |  |  |
| Unadjusted | 1.21 | 1.11 | 1.31 | <0.001 |
| Adjusted | 1.02 | 0.94 | 1.12 | 0.587 |
| <u>Healthcare disruption type – unadjusted</u> |  |  |  |  |
| Appointments | 1.23 | 1.12 | 1.35 | <0.001 |
| Medications | 0.95 | 0.77 | 1.18 | 0.640 |
| Procedures | 1.05 | 0.94 | 1.19 | 0.374 |
| <u>Healthcare disruption type - adjusted</u> |  |  |  |  |
| Appointments | 1.08 | 0.98 | 1.19 | 0.138 |
| Medications | 1.10 | 0.91 | 1.33 | 0.323 |
| Procedures | 0.95 | 0.85 | 1.07 | 0.377 |

Note: Estimates are Odds Ratios. CI = Confidence Interval. Model adjustment includes the following variables: age, sex, ethnicity, housing tenure, whether had COVID-19, self-rated health status, and longitudinal cohort).

## Discussion

### Key results

Our study presents the first empirical investigation utilising linked individual-level data to examine the impacts of healthcare disruption on avoidable hospitalisations. We estimated that 35% of people in England experienced disrupted access to healthcare, with disruption to appointments (e.g., seeing a GP or healthcare professional) being most common. Overall, individuals who reported any form of disruption in accessing healthcare were more likely to have been admitted to hospital for an avoidable or potentially preventable condition between 1^st^ March 2020 and 25^th^ August 2022. We found an 80%, 68% and 93% increase in risk of hospitalisation for any, acute and chronic ambulatory care sensitive conditions respectively. Individuals who reported disruption in accessing medications or ambulatory appointments were both positively associated to people being more likely to be hospitalised for any ambulatory care sensitive conditions. Individuals who reported disruption in accessing procedures were more likely to be hospitalised for chronic ambulatory care sensitive or emergency urgent care sensitive conditions. Finally, we found no associations to vaccination status following adjustment as a falsification test.

### Interpretation

Untangling the consequences of healthcare disruption is difficult due to the short- and longer-term implications of missed diagnostic tests, preventative treatments and anxiety from accessing healthcare (14). Our study presents novel evidence that people whose access to healthcare was disrupted were more likely to have an avoidable or potentially preventable hospitalisation, at least in the short term (∼two years post pandemic onset). Similarly, we found that disruption to specific parts of the health system, including to appointments or procedures, were also associated with greater risk of different types of avoidable hospitalisations.

There are several potential explanations for these associations. Appointments with healthcare professional provide people with opportunities to seek advice, access secondary care, receive diagnostic tests, and receive treatment. Disruptions may delay care that is needed, with people needing hospitalisation as diseases progress (for example, presenting at later disease stages that are harder to treat). In particular, sudden changes in health often prompt people to seek a consultation, which may explain why we found an association with acute ambulatory care sensitive conditions. Similarly, disruptions to procedures (e.g., surgery, treatment) may lead to exacerbations of existing and longer-term conditions, or disease progression that would otherwise have been treated, which may produce avoidable hospital admissions (especially when associated with chronic ambulatory care sensitive conditions). Future research should tease out these specific pathways where disruption leads to an avoidable hospitalisation to identify mechanisms that could mitigate the effects of disruption of services.

Few people experienced problems with obtaining medications (6%), which were also rarely associated with avoidable hospitalisations. Our findings may suggest that medicines supply was resilient during the pandemic. Even during periods of greatest disruption, when there were lockdowns, pharmacies were deemed essential services and remained open in the UK. While they did experience some issues with staff sickness and medicine shortages (23), they adapted successfully, aided by remote GP consultations and home deliveries (2).

Our findings demonstrate the need to increase investment in the health system to counter the negative effects of healthcare disruption resulting from the COVID-19 pandemic. While NHS activity has returned to some extent, it has not returned to 2019 levels (24) and the NHS has struggled to clear the backlog of treatments, diagnostic tests, procedures and appointments (25). More recent disruption during winter 2022-23, with low rates of staff retention, chronic underfunding, healthcare worker strikes, high levels of staff illness, high prevalence of influenza and COVID-19, and persisting waiting lists have compounded the pandemic-related disruption (13,25). However, UK’s dire economic situation following Brexit and the catastrophic mini-budget in 2022, coupled with a lack of political will to increase funding, will make it difficult for the NHS to tackle its legacy of underinvestment in labour and capital (3).

Our study has several strengths. We combined data from seven individual-level longitudinal studies which are drawn from independently nationally representative samples (although this may not translate to a representative pooled sample entirely). Using only a single study would have restricted our sample size for events or limited analyses to certain demographic groups. The ability to systematically link self-reported data on disruption experienced to participant’s electronic health records within the UK Longitudinal Linkage Collaboration has been crucial to overcome previous barriers of linking survey data with health records (26). By combining individual-level longitudinal studies and electronic health records we complement their individual strengths (i.e., depth of context about individuals alongside objective hospital admissions data). Importantly, our results contrast with research using electronic health records alone, which showed falls in avoidable hospitalisations during periods of greatest disruption (24). These different insights demonstrate how analysis of population level routine records can be misleading where they don’t have the same level of detail linked individual-level survey data often contain.

### Limitations

Our analyses are observational and have limited ability to draw causal inferences. We are unable to link specific experiences of disruption to particular adverse events, and not all avoidable hospitalisations will be due to the disruption of care. Our exposure measures were self-reported and may be subject to biases. For example, “plaintive set” can influence self-reported measures and may have inflated reports of exposure to disruption. In the situation where this reporting bias also influences outcome measures in the same direction biased associations can arise. Hospital admission depends on an individual presenting to a health facility and complaining of their condition. We have shown elsewhere that this can lead to bias (27), although this may have been less of a risk during our study given the general reluctance of hospitals to admit patients during the study period (24).

Inconsistency of measures across cohorts limited our ability to control for potential confounders. There may be some bias introduced by data linkage due to incomplete or incorrect matching (26,28), which may disproportionately impact on marginalised groups and people who may have migrated to other parts of the UK. Biases may have been introduced where study participants did not consent to linkage to their health records (28), although the impacts on our exposures was limited (Appendix Table A). Due to the rarity of our outcome variables, we were unable to explore whether experiences of healthcare disruption were greater in particular subgroups or demographics. This is pertinent since the topics we examine are not evenly felt across the UK. For example, healthcare disruption was disproportionately experienced in socioeconomically deprived communities (3,7,14), with avoidable hospitalisations also higher in deprived areas (29). The rarity of our outcomes produces wide confidence intervals so that, even with a large combined sample, power is limited.

Unplanned hospitalisations may only occur after a long period, stretching beyond our study period (14). Thus, we may have under-estimated the impacts of healthcare disruption. It will be important to follow experiences of our cohort members over longer time periods to determine whether this is the case. Additionally, some have questioned how sensitive avoidable hospitalisations are to health system performance, as they may be affected by issues beyond the control of health systems as well (e.g., socioeconomic deprivation) (17,30). Future research should investigate other outcomes, including moving away from composite indicators, to understand the pathways through which disruption impacts individuals. The current expansion of the UK LLC to include additional longitudinal studies and a larger participant sample size may facilitate this.

The use of vaccination status as a falsification test is a strength, but cannot definitively confirm a lack of residual confounding (21,22). People who were not vaccinated may be less connected to the health system and therefore less likely to experience disruption. Future research may consider more robust indicators.

### Conclusions

The external shock to the health system caused by the COVID-19 pandemic significantly disrupted access to healthcare and this impact is having negative impacts of hospital admissions that could be potentially preventable. As narratives on how to respond to a pandemic, continued disruption to the NHS, and how to ‘build back better’ develop, our paper highlights the need to increase healthcare investment to tackle the short- and long-term implications of the pandemic.

## Data Availability

The datasets used in the study are all safeguarded and secured data and therefore cannot be openly shared. All data can be accessed by accredited researchers via application to the UK Longitudinal Linkage Collaboration (https://ukllc.ac.uk/). All analytical code to replicate analyses can be found at https://github.com/markagreen/healthcare_disruption_LLC.

https://github.com/markagreen/healthcare_disruption_LLC

## Acknowledgements

This work was supported by the Medical Research Council [grant numbers MR/W021242/1, MC_UU_00022/2], NHS Research Scotland [SCAF/15/02] and the Scottish Government Chief Scientist Office [SPHSU17]. RJS is funded by Health Data Research UK (SS005). JM is partly funded by the National Institute for Health and Care Research Applied Research Collaboration West (NIHR ARC West). The funders played no role in the design of the study, analyses, write up or plan to submit the paper for publication.

The full statement, listing the names of all relevant NCS Consortium staff, can be found here: https://www.ucl.ac.uk/covid-19-longitudinal-health-wellbeing/convalescence-studycollaborative.

We thank the NHS and particularly NHS Digital for their work in curating participants’ health records and for making these available for public benefit research designed to improve health services. We thank the University of Leicester for providing geospatial data and Ordnance Survey for providing AddressBase® Plus.

This project has been approved by UK LLC and its contributing data owners and information on this project and its outputs can be accessed via UK LLC’s website (Data Use Register | UK Longitudinal Linkage Collaboration; https://ukllc.ac.uk/) and UK LLC’s GitHub (UK Longitudinal Linkage Collaboration GitHub; https://github.com/UKLLC). The UK LLC has ethical approval from the Health Research Authority Research Ethics Committee to support COVID-19 research (Haydock Committee; ref: 20/NW/0446).

UK LLC is a Trusted Research Environment developed and operated by the Universities of Bristol and Edinburgh using an underlying ‘Secure eResearch Platform’ infrastructure (https://serp.ac.uk/) provided by Swansea University for longitudinal research. The UK LLC is an initiative of the UKRI-funded Longitudinal Health and Wellbeing National Core Study led by University College London (Grant code: MC_PC_20059). This work uses data accessed within UK LLC’s Trusted Research Environment (TRE), hosted by the Secure eResearch Platform (SeRP UK). We thank the SeRP UK Team at Swansea University and NHS Digital Health and Care Wales for providing the TRE’s infrastructure and support.

This work uses data provided by participants of the contributing longitudinal population studies (LPS) within the UK LLC TRE, which have been collected through their longitudinal study or as part of their care and support and/or interactions with UK government services. We wish to recognise and thank the study participants and each contributing LPS team, including data managers, administrators and those collecting data. We thank the following LPS for contributing data that made this research possible:

- 1970 British Cohort Study (BCS70)
- English Longitudinal Study of Ageing (ELSA)
- Medical Research Council (MRC) National Survey of Health and Development (NSHD)
- Millennium Cohort Study (MCS)
- National Child Development Study (NCDS)
- Next Steps
- The UK Household Longitudinal Study (Understanding Society).

A full list of acknowledgments, including support for each study, is provided in the supplementary materials.

