## Appendix for "Associations between reported healthcare disruption due to COVID-19 and avoidable hospitalisation: Evidence from seven linked longitudinal studies for England"

**Table A: Differences in experiences of healthcare disruption between individuals who could be linked to NHS Digital Hospital Episode Statistics.**

| **Variable (disruption to…)** | **Linked (i.e., data used in analyses)** | **Did not consent to linkage or linkage was not possible** |
| --- | --- | --- |
| Appointments | 25.0 | 23.3 |
| Medications | 4.9 | 4.6 |
| Procedures | 18.1 | 15.5 |

**Table B: Analytical sample size by cohort.**

| Cohort | Sample size | Percentage |
| --- | --- | --- |
| 1970 Birth Cohort | 4122 | 14.1 |
| English Longitudinal Study of Ageing | 5567 | 19.0 |
| Millennium Cohort Study | 3350 | 11.4 |
| 1958 Birth Cohort | 4725 | 16.1 |
| NextSteps | 2891 | 9.9 |
| 1946 Birth Cohort | 1813 | 6.2 |
| Understanding Society | 6808 | 23.3 |
| Total | 29276 | 100 |

**Table C: Description of healthcare disruption questions used to measure each exposure (after Maddock et al. 2021).**

| **Cohort** | **Medication** | **Appointments** | **Procedures** |
| --- | --- | --- | --- |
| National Survey of Health and Development  National Child Development Study  1970 British Cohort Study  Next Steps  Millennium Cohort Study | Since the Coronavirus outbreak in March, have you had any difficulty obtaining any of your prescribed medication? | Q1: At the time of the Coronavirus outbreak in March, did you have an in-patient or out-patient appointment booked at a hospital for a consultation, investigation, treatment or surgery?  Q2: Have you now had your in/ out-patient hospital appointment for a consultation, investigation or treatment?  Q3: Did your (last) appointment take place on the planned date or was it delayed?  Q4: Why has your in-/out-patient hospital appointment for a consultation, investigation or treatment not taken place? | Q1: At the time of the Coronavirus outbreak in March, did you have an in-patient or out-patient appointment booked at a hospital for a consultation, investigation, treatment or surgery?  Q2: Have you now had your surgery?  Q3: Did your (last) surgery take place on the planned date or was it delayed?  Q4: Why has your surgery not taken place? |
| English Longitudinal Study of Ageing | Since the coronavirus outbreak, have you been able to get access to your regular medications? | Q1: Since the coronavirus outbreak, have you wanted to see or talk to a GP?  Q2: Have you been able to see or talk to a GP? | Since the coronavirus outbreak, have you had a hospital operation or treatment cancelled? |
| Understanding Society | Q1: Still thinking about your situation now, have you been able to access the NHS services you need: Prescription medicine?  Q2: Still thinking about your situation now, have you been able to access the community health and social care services and support you need... Over the counter medications? | Thinking about your situation now, have you been able to access the NHS services you need to help manage your condition(s) over the last 4 weeks?  Q1: GP or primary care practice staff?  Q2: Hospital or clinic outpatient?  Q3: Hospital or clinic inpatient?  Q4: [since previous survey] have you had or been waiting for NHS treatment? Please select all that apply. | Q1: [since previous survey] have you had or been waiting for NHS treatment? Please select all that apply.  Q2: Has your treatment plan(s) been changed in any way? |

**Table D: Missing data in pooled analytical sample across outcome, exposure and control variables.**

| Variable | Frequency | Percentage |
| --- | --- | --- |
| Age | 0 | 0.0 |
| Sex | 63 | 0.2 |
| Ethnicity | 994 | 3.4 |
| Housing tenure | 3349 | 11.4 |
| Self-rated health | 463 | 1.6 |
| Index of multiple deprivation | 193 | 0.7 |
| Disruption to appointments | 1460 | 5.0 |
| Disruption to medications | 8919 | 30.5 |
| Disruption to procedures | 5513 | 18.8 |

**Table E: Descriptive statistics for control variables adjusted for in analyses (note: value is mean for age and percentages (%) for all other variables).**

| Measure | Frequency | Unweighted value | Weighted value |
| --- | --- | --- | --- |
| Age (mean) |  | 52.9 | 52.9 |
| Female (%) | 15722 | 53.7 | 50.1 |
| Male (%) | 13554 | 46.3 | 49.9 |
| White (%) | 26746 | 91.4 | 92.4 |
| Not White (%) | 2530 | 8.6 | 7.6 |
| Own home (%) | 14667 | 50.1 | 50.2 |
| Do not own home (%) | 14609 | 49.9 | 49.8 |
| Good health (%) | 23079 | 78.8 | 75.8 |
| Poor health (%) | 6197 | 21.2 | 24.2 |
| IMD Quintile 1 (%) - Most Deprived | 3631 | 12.4 | 16.3 |
| IMD Quintile 2 (%) | 4774 | 16.3 | 17.9 |
| IMD Quintile 3 (%) | 6054 | 20.7 | 20.1 |
| IMD Quintile 4 (%) | 7144 | 24.4 | 22.3 |
| IMD Quintile 5 (%) - Least Deprived | 7673 | 26.2 | 23.3 |

**Table F: Summary of outcome variables across two sensitivity analyses.**

| Measure | 1: Include events post survey (events) | | | 2: Survival analysis (days) | |
| --- | --- | --- | --- | --- | --- |
|  | Frequency | Percent | Weighted Percent | Unweighted Mean | Weighted Mean |
| Total admissions | 2462 | 8.41 | 9.30 | 204.82 | 198.82 |
| Ambulatory care sensitive any | 503 | 1.72 | 2.44 | 216.81 | 215.08 |
| Ambulatory care sensitive acute | 218 | 0.74 | 0.90 | 218.82 | 223.93 |
| Ambulatory care sensitive chronic | 240 | 0.82 | 0.95 | 211.79 | 192.61 |
| Ambulatory care sensitive vaccine preventable | 60 | 0.20 | 0.67 | 239.22 | 241.43 |
| Emergency urgent care sensitive | 371 | 1.27 | 1.42 | 220.46 | 215.87 |

**Table G: Logistic regression results for a sensitivity analysis excluding outcome events that occurred before the last known survey date to examine the association between experiences of healthcare disruption to avoidable hospitalisations (i.e., results presented in Table 2).**

| **Model** | **Odds Ratio** | **Lower 95% CI** | **Higher 95% CI** | **P value** |
| --- | --- | --- | --- | --- |
| Any ambulatory care sensitive | | |  |  |
| Unadjusted | 2.72 | 1.49 | 4.95 | 0.001 |
| Adjusted | 1.70 | 1.18 | 2.46 | 0.005 |
| Acute ambulatory care sensitive | | |  |  |
| Unadjusted | 1.60 | 0.99 | 2.60 | 0.056 |
| Adjusted | 1.45 | 0.86 | 2.44 | 0.161 |
| Chronic ambulatory care sensitive | | |  |  |
| Unadjusted | 3.06 | 2.06 | 4.54 | <0.001 |
| Adjusted | 1.93 | 1.32 | 2.84 | 0.001 |
| Vaccine-preventable ambulatory care sensitive | | | |  |
| Unadjusted | 4.61 | 0.82 | 25.89 | 0.083 |
| Adjusted | 1.35 | 0.64 | 2.83 | 0.428 |
| Emergency urgent care sensitive | | |  |  |
| Unadjusted | 1.49 | 1.05 | 2.11 | 0.024 |
| Adjusted | 1.09 | 0.77 | 1.53 | 0.629 |
| Any hospital admission | |  |  |  |
| Unadjusted | 2.32 | 1.97 | 2.73 | <0.001 |
| Adjusted | 1.68 | 1.41 | 2.00 | <0.001 |

**Table H: Cox regression results for a sensitivity analysis for time to hospital admission (month) of events happening after the last known survey date to examine the association between experiences of healthcare disruption to avoidable hospitalisations (i.e., results presented in Table 2).**

| **Model** | **Hazards Ratio** | **Lower 95% CI** | **Higher 95% CI** | **P value** |
| --- | --- | --- | --- | --- |
| Any ambulatory care sensitive | | |  |  |
| Unadjusted | 2.16 | 1.81 | 2.58 | <0.001 |
| Adjusted | 1.51 | 1.26 | 1.82 | <0.001 |
| Acute ambulatory care sensitive | | |  |  |
| Unadjusted | 1.90 | 1.46 | 2.48 | <0.001 |
| Adjusted | 1.67 | 1.26 | 2.20 | <0.001 |
| Chronic ambulatory care sensitive | | |  |  |
| Unadjusted | 2.46 | 1.91 | 3.17 | <0.001 |
| Adjusted | 1.53 | 1.17 | 2.00 | 0.002 |
| Vaccine-preventable ambulatory care sensitive | | | |  |
| Unadjusted | 2.01 | 1.21 | 3.33 | 0.007 |
| Adjusted | 1.00 | 0.59 | 1.69 | 0.987 |
| Emergency urgent care sensitive | | |  |  |
| Unadjusted | 1.84 | 1.50 | 2.25 | <0.001 |
| Adjusted | 1.32 | 1.07 | 1.63 | 0.011 |
| Any hospital admission | |  |  |  |
| Unadjusted | 2.43 | 2.24 | 2.63 | <0.001 |
| Adjusted | 1.73 | 1.59 | 1.88 | <0.001 |

**Table I: Logistic regression results for a sensitivity analysis excluding outcome events that occurred before the last known survey date to examine the association between experiences of healthcare disruption to avoidable hospitalisations (i.e., results presented in Table 3).**

| **Model** | **Odds Ratio** | **Lower 95% CI** | **Higher 95% CI** | **P value** |
| --- | --- | --- | --- | --- |
| Any ambulatory care sensitive - unadjusted model | | | | |
| Appointments | 2.12 | 0.91 | 4.90 | 0.081 |
| Medications | 3.90 | 1.03 | 14.73 | 0.046 |
| Procedures | 1.12 | 0.44 | 2.86 | 0.822 |
| Any ambulatory care sensitive - adjusted model | | | |  |
| Appointments | 1.38 | 0.90 | 2.10 | 0.150 |
| Medications | 3.42 | 1.45 | 8.00 | 0.005 |
| Procedures | 1.20 | 0.73 | 1.95 | 0.489 |
| Acute ambulatory care sensitive - unadjusted model | | | |  |
| Appointments | 1.17 | 0.73 | 1.86 | 0.503 |
| Medications | 1.17 | 0.30 | 4.62 | 0.818 |
| Procedures | 1.55 | 1.01 | 2.41 | 0.047 |
| Acute ambulatory care sensitive - adjusted model | | | |  |
| Appointments | 1.04 | 0.66 | 1.65 | 0.863 |
| Medications | 1.17 | 0.26 | 5.31 | 0.840 |
| Procedures | 1.63 | 1.08 | 2.48 | 0.021 |
| Chronic ambulatory care sensitive - unadjusted model | | | |  |
| Appointments | 1.45 | 0.76 | 2.75 | 0.255 |
| Medications | 0.84 | 0.44 | 1.63 | 0.622 |
| Procedures | 2.94 | 1.51 | 5.70 | 0.002 |
| Chronic ambulatory care sensitive - adjusted model | | | |  |
| Appointments | 1.20 | 0.68 | 2.08 | 0.532 |
| Medications | 0.89 | 0.39 | 2.01 | 0.777 |
| Procedures | 2.27 | 1.26 | 4.10 | 0.006 |
| Vaccine-preventable ambulatory care sensitive - unadjusted model | | | | |
| Appointments | 5.58 | 1.16 | 26.84 | 0.032 |
| Medications |  |  |  |  |
| Procedures | 0.15 | 0.02 | 0.96 | 0.045 |
| Vaccine-preventable ambulatory care sensitive - adjusted model | | | | |
| Appointments | 1.45 | 0.79 | 2.66 | 0.234 |
| Medications |  |  |  |  |
| Procedures | 0.28 | 0.11 | 0.72 | 0.008 |
| Emergency urgent care sensitive - unadjusted model | | | |  |
| Appointments | 0.95 | 0.60 | 1.52 | 0.841 |
| Medications | 0.87 | 0.50 | 1.54 | 0.632 |
| Procedures | 2.08 | 1.22 | 3.53 | 0.007 |
| Emergency urgent care sensitive - adjusted model | | | |  |
| Appointments | 0.84 | 0.55 | 1.30 | 0.428 |
| Medications | 0.87 | 0.44 | 1.68 | 0.672 |
| Procedures | 1.72 | 1.06 | 2.75 | 0.026 |
| Any hospital admission - unadjusted model | | | |  |
| Appointments | 1.32 | 1.05 | 1.67 | 0.018 |
| Medications | 1.01 | 0.68 | 1.51 | 0.963 |
| Procedures | 2.14 | 1.67 | 2.75 | 0.000 |
| Any hospital admission - adjusted model | | | |  |
| Appointments | 1.14 | 0.90 | 1.43 | 0.274 |
| Medications | 1.14 | 0.67 | 1.92 | 0.638 |
| Procedures | 1.84 | 1.43 | 2.34 | <0.001 |

Note: Results for vaccine-preventable ambulatory care sensitive conditions and disruption to medications were not robust due to small number issues.

**Table J: Cox regression results for a sensitivity analysis excluding outcome events that occurred before the last known survey date to examine the association between experiences of healthcare disruption to avoidable hospitalisations (i.e., results presented in Table 3).**

| **Model** | **Hazards Ratio** | **Lower 95% CI** | **Higher 95% CI** | **P value** |
| --- | --- | --- | --- | --- |
| Any ambulatory care sensitive - unadjusted model | | | | |
| Appointments | 1.54 | 1.39 | 1.70 | <0.001 |
| Medications | 0.93 | 0.79 | 1.11 | 0.433 |
| Procedures | 2.03 | 1.84 | 2.25 | <0.001 |
| Any ambulatory care sensitive - adjusted model | | | |  |
| Appointments | 1.28 | 1.17 | 1.40 | <0.001 |
| Medications | 1.25 | 1.05 | 1.48 | 0.010 |
| Procedures | 1.70 | 1.55 | 1.88 | <0.001 |
| Acute ambulatory care sensitive - unadjusted model | | | |  |
| Appointments | 1.54 | 1.39 | 1.70 | <0.001 |
| Medications | 0.93 | 0.79 | 1.11 | 0.431 |
| Procedures | 2.03 | 1.84 | 2.25 | <0.001 |
| Acute ambulatory care sensitive - adjusted model | | | |  |
| Appointments | 1.28 | 1.17 | 1.40 | <0.001 |
| Medications | 1.23 | 1.04 | 1.46 | 0.014 |
| Procedures | 1.70 | 1.54 | 1.86 | <0.001 |
| Chronic ambulatory care sensitive - unadjusted model | | | |  |
| Appointments | 1.54 | 1.39 | 1.70 | <0.001 |
| Medications | 0.93 | 0.79 | 1.11 | 0.432 |
| Procedures | 2.03 | 1.84 | 2.25 | <0.001 |
| Chronic ambulatory care sensitive - adjusted model | | | |  |
| Appointments | 1.28 | 1.17 | 1.40 | <0.001 |
| Medications | 1.25 | 1.05 | 1.48 | 0.011 |
| Procedures | 1.70 | 1.55 | 1.88 | <0.001 |
| Vaccine-preventable ambulatory care sensitive - unadjusted model | | | | |
| Appointments | 1.52 | 1.39 | 1.68 | <0.001 |
| Medications |  |  |  |  |
| Procedures | 2.03 | 1.84 | 2.25 | <0.001 |
| Vaccine-preventable ambulatory care sensitive - adjusted model | | | | |
| Appointments | 1.27 | 1.16 | 1.40 | <0.001 |
| Medications |  |  |  |  |
| Procedures | 1.70 | 1.54 | 1.86 | <0.001 |
| Emergency urgent care sensitive - unadjusted model | | | |  |
| Appointments | 1.54 | 1.39 | 1.68 | <0.001 |
| Medications | 0.94 | 0.79 | 1.11 | 0.447 |
| Procedures | 2.03 | 1.84 | 2.25 | <0.001 |
| Emergency urgent care sensitive - adjusted model | | | |  |
| Appointments | 1.28 | 1.16 | 1.40 | <0.001 |
| Medications | 1.25 | 1.05 | 1.49 | 0.010 |
| Procedures | 1.70 | 1.55 | 1.88 | <0.001 |
| Any hospital admission - unadjusted model | | | |  |
| Appointments | 1.54 | 1.39 | 1.68 | <0.001 |
| Medications | 0.94 | 0.79 | 1.12 | 0.471 |
| Procedures | 2.03 | 1.84 | 2.25 | <0.001 |
| Any hospital admission - adjusted model | | | |  |
| Appointments | 1.28 | 1.17 | 1.40 | <0.001 |
| Medications | 1.25 | 1.05 | 1.48 | 0.012 |
| Procedures | 1.72 | 1.55 | 1.88 | <0.001 |

Note: Results for vaccine-preventable ambulatory care sensitive conditions and disruption to medications were not robust due to small number issues.

**Table K: Detailed information of the seven longitudinal population studies.**

| **BCS70: 1970 British Cohort Study** | |
| --- | --- |
| **Description of Study Population** (including citations and references if required) | The 1970 British Cohort Study (BCS70) follows the lives of more than 17,000 people born in England, Scotland and Wales in a single week of 1970. Over the course of cohort members’ lives, BCS70 has collected information on health, physical, educational and social development, and economic circumstances, among other factors.  Since the birth survey in 1970, there have been nine ‘sweeps’ of all cohort members at ages 5, 10, 16, 26, 30, 34, 38, 42 and most recently at 46 (a biomedical data collection).  The Age 51 Sweep is currently in the field (2022).  Data have been collected from a number of different sources, including the midwife present at birth, parents of the cohort members, head and class teachers, school health service personnel and the cohort members themselves.  The data have been collected in a variety of ways, including via paper and electronic questionnaires, clinical records, medical examinations, biological samples, physical measurements, tests of ability, educational assessments and diaries.  The study is conducted by the Centre for Longitudinal Studies. |
| **Acknowledgements** | BCS70 is core-funded by the ESRC. |
| **Ethics** | Ethics approval has been obtained for each follow-up from an NHS Research Ethics Committee (REC) since 2000. In addition, separate REC approval is in place to cover the ongoing activities of the study in between major sweeps of data collection (i.e. Keeping in touch with and tracing cohort members; cleaning, documenting and providing access to the data for research; and linking data from administrative sources to survey data to increase the utility of the data for research). |
| **Website for Data Requests** | <https://cls.ucl.ac.uk/cls-studies/bcs70/> |
| **COVID-19 survey dates** | Three surveys: May 2020, Sept-Oct 2020 and Feb-Mar 2021 |
| **Citation** | University College London, UCL Institute of Education, Centre for Longitudinal Studies. (2022). COVID-19 Survey in Five National Longitudinal Cohort Studies: Millennium Cohort Study, Next Steps, 1970 British Cohort Study and 1958 National Child Development Study, 2020-2021. [data collection]. 4th Edition. UK Data Service. SN: 8658, DOI: 10.5255/UKDA-SN-8658-4 |

| **ELSA: English Longitudinal Study of Ageing** | |
| --- | --- |
| **Description of Study Population** (including citations and references if required) | The English Longitudinal Study of Ageing (ELSA) is a unique and rich resource of information on the dynamics of health, social, wellbeing and economic circumstances in the English population aged 50 and older ^1^.  The original sample was drawn from households that had previously responded to the Health Survey for England (HSE) between 1998 and 2001. The main fieldwork began in March 2002. The same group of respondents have been interviewed at two-yearly interviews .  ^1^Banks J, Batty GD, Breedvelt JJF, Coughlin K, Crawford R, Marmot M, Nazroo J, Oldfield Z, Steel N, Steptoe A, Wood M, Zaninotto P (2021) English Longitudinal Study of Ageing: Waves 0-9, 1998-2019 |
| **Acknowledgements** | The English Longitudinal Study of Ageing was developed by a team of researchers based at University College London, NatCen Social Research, the Institute for Fiscal Studies, the University of Manchester and the University of East Anglia. The data were collected by NatCen Social Research. The funding is currently provided by the National Institute on Aging (Ref: R01AG017644) and by a consortium of UK government departments: Department for Health and Social Care; Department for Transport; Department for Work and Pensions, which is coordinated by the National Institute for Health Research (NIHR, Ref: 198-1074). Funding has also been provided by the Economic and Social Research Council (ESRC). |
| **Ethics** | <https://www.elsa-project.ac.uk/ethical-approval> |
| **Website for Data Requests** | <https://www.elsa-project.ac.uk/data-and-documentation> |
| **COVID-19 survey dates** | Two surveys: Jun-Jul 2020 and Nov-Dec 2020 |
| **Citation** | Marmot, M., Pacchiotti, B., Banks, J., Steel, N., Oldfield, Z., Nazroo, J., Dangerfield, P., Coughlin, K., Zaninotto, P., Crawford, R., Steptoe, A., Addario, G., Wood, M., Batty, G. David. (2022). English Longitudinal Study of Ageing COVID-19 Study, Waves 1-2, 2020. [data collection]. 3rd Edition. UK Data Service. SN: 8688, DOI: 10.5255/UKDA-SN-8688-3 |

|  | **MCS: Millennium Cohort Study** | | |
| --- | --- | --- | --- |
|  | **Description of Study Population** (including citations and references if required) | | The Millennium Cohort Study (MCS) is following the lives of young people born across England, Scotland, Wales and Northern Ireland in 2000-02. The study began with an original sample of 18,818 cohort members. The study is designed and led by the Centre for Longitudinal Studies (CLS) at University College London.  The broad aim of the study is to examine the impact that circumstances and experiences at one stage of life have on outcomes and achievements in later life. Since the baseline survey at age 9 months, there have been six major ‘sweeps’ at ages 3, 5, 7, 11, 14 and 17. The next sweep, at age 22, is currently under development.  Data have been collected from a number of different sources, including the cohort members and their parents and teachers. The data have been collected in a variety of ways, including via paper and electronic questionnaires, biological samples, physical measurements, tests of ability, and linked educational attainment and health records.  The information collected forms a high quality data resource for scientific investigations across a full range of domains of individuals’ lives and across different points in time in them. The study has been designed to ensure comparability with other major cohort studies both in the UK and internationally and to permit the examination of links between social change and the changing experiences of different cohorts.  <https://www.llcsjournal.org/index.php/llcs/article/view/410/0>  <https://academic.oup.com/ije/article/43/6/1719/703283> |
|  | **Acknowledgements** | | MCS is core-funded by the ESRC and co-funded by a consortium of government departments. |
|  | **Ethics** | | Ethics approval has been obtained for each follow-up from an NHS Research Ethics Committee (REC). In addition, separate REC approval is in place to cover the ongoing activities of the study in between major sweeps of data collection (i.e. Keeping in touch with and tracing cohort Members; cleaning, documenting and providing access to the data for research and linking data from administrative sources to survey data to increase the utility of the data for research. |
|  | **Website for Data Requests** | | <https://cls.ucl.ac.uk/cls-studies/mcs/> |
|  | **COVID-19 survey dates** | | Three surveys: May 2020, Sept-Oct 2020 and Feb-Mar 2021 |
|  | **Citation** | | University College London, UCL Institute of Education, Centre for Longitudinal Studies. (2022). COVID-19 Survey in Five National Longitudinal Cohort Studies: Millennium Cohort Study, Next Steps, 1970 British Cohort Study and 1958 National Child Development Study, 2020-2021. [data collection]. 4th Edition. UK Data Service. SN: 8658, DOI: 10.5255/UKDA-SN-8658-4 |
| **NCDS: National Child Development Study** | | | |
| **Description of Study Population** (including citations and references if required) | | The National Child Development Study (NCDS) is a continuing longitudinal study that seeks to follow the lives of all those living in Great Britain who were born in one particular week in 1958. Conducted by the Centre for Longitudinal Studies , the aim of the study is to improve understanding of the factors affecting human development over the whole lifespan. It collects information on physical and educational development, economic circumstances, employment, family life, health behaviour, wellbeing, social participation and attitudes.  The broad aim of the study is to examine the impact that circumstances and experiences at one stage of life have on outcomes and achievements in later life. Since the birth survey in 1958, there have been ten ‘sweeps’ of all cohort members at ages 7, 11, 16, 23, 33, 42, 44/5 (a biomedical collection) 46, 50 and most recently at 55. The Age 62 Sweep is currently in the field (2022).  Data have been collected from a number of different sources, including the midwife present at birth, parents of the cohort members, teachers, doctors and the cohort members themselves. The data have been collected in a variety of ways, including via paper and electronic questionnaires, clinical records, medical examinations, biological samples, physical measurements, tests of ability and educational assessments.  The information collected forms a high quality data resource for scientific investigations across a full range of domains of individuals’ lives and across different points in time in them. The study has been designed to ensure comparability with other major cohort studies and to permit the examination of links between social change and the changing experiences of different cohorts.  <https://cls.ucl.ac.uk/cls-studies/1958-national-child-development-study/> | |
| **Acknowledgements** | | NCDS is core-funded by the ESRC. | |
| **Ethics** | | Ethics approval has been obtained for each follow-up from an NHS Research Ethics Committee (REC) since 2000. In addition, separate REC approval is in place to cover the ongoing activities of the study in between major sweeps of data collection (i.e. Keeping in touch with and tracing cohort members; cleaning, documenting and providing access to the data for research; and linking data from administrative sources to survey data to increase the utility of the data for research). | |
| **Website for Data Requests** | | <https://cls.ucl.ac.uk/cls-studies/ncds/> | |
| **COVID-19 survey dates** | | Three surveys: May 2020, Sept-Oct 2020 and Feb-Mar 2021 | |
| **Citation** | | University College London, UCL Institute of Education, Centre for Longitudinal Studies. (2022). COVID-19 Survey in Five National Longitudinal Cohort Studies: Millennium Cohort Study, Next Steps, 1970 British Cohort Study and 1958 National Child Development Study, 2020-2021. [data collection]. 4th Edition. UK Data Service. SN: 8658, DOI: 10.5255/UKDA-SN-8658-4 | |

| **Next Steps** | |
| --- | --- |
| **Description of Study Population** (including citations and references if required) | Next Steps (previously known as the Longitudinal Study of Young People in England (LSYPE1)) is a major longitudinal study that follows the lives of around 16,000 people born in 1989-90. The first seven sweeps of the study (2004-2010) were funded and managed by the Department for Education and mainly focused on the educational and early labour market experiences of young people.  The study began in 2004 and included young people in Year 9 who attended state and independent schools in England. Following the initial survey at age 13-14, the cohort members were interviewed every year until 2010.  In 2013 the management of Next Steps was transferred to the Centre for Longitudinal Studies (CLS) at the IOE, UCL’s Faculty of Education and Society. The first sweep conducted by CLS aimed to find out how the lives of the cohort members had turned out at age 25. It maintained the strong focus on education, but the content was broadened to become a more multi-disciplinary research resource.  The Age 32 Sweep is currently in the field (2022).  [https://doc.ukdataservice.ac.uk/doc/5545/mrdoc/pdf/next_steps_userguide_to_the_redeposit_of_sweeps_1to7_may2020.pdf](https://eur01.safelinks.protection.outlook.com/?url=https%3A%2F%2Fdoc.ukdataservice.ac.uk%2Fdoc%2F5545%2Fmrdoc%2Fpdf%2Fnext_steps_userguide_to_the_redeposit_of_sweeps_1to7_may2020.pdf&data=05%7C01%7Cmorag.henderson%40ucl.ac.uk%7Ca5279433ef414feb792808da6319282f%7C1faf88fea9984c5b93c9210a11d9a5c2%7C0%7C0%7C637931256931551677%7CUnknown%7CTWFpbGZsb3d8eyJWIjoiMC4wLjAwMDAiLCJQIjoiV2luMzIiLCJBTiI6Ik1haWwiLCJXVCI6Mn0%3D%7C3000%7C%7C%7C&sdata=ColBf1sx2sz8g95l4WKpe3z%2B1ymQyhVgJOvHoubF3BY%3D&reserved=0)  [https://doc.ukdataservice.ac.uk/doc/5545/mrdoc/pdf/nextsteps_age25_survey_user_guide_v3.pdf](https://eur01.safelinks.protection.outlook.com/?url=https%3A%2F%2Fdoc.ukdataservice.ac.uk%2Fdoc%2F5545%2Fmrdoc%2Fpdf%2Fnextsteps_age25_survey_user_guide_v3.pdf&data=05%7C01%7Cmorag.henderson%40ucl.ac.uk%7Ca5279433ef414feb792808da6319282f%7C1faf88fea9984c5b93c9210a11d9a5c2%7C0%7C0%7C637931256931551677%7CUnknown%7CTWFpbGZsb3d8eyJWIjoiMC4wLjAwMDAiLCJQIjoiV2luMzIiLCJBTiI6Ik1haWwiLCJXVCI6Mn0%3D%7C3000%7C%7C%7C&sdata=Xy8K4SuQRsqAMbqs19kgBiff4EQvnlHm6HANwXQY%2B20%3D&reserved=0)  <https://cls.ucl.ac.uk/cls-studies/next-steps/> |
| **Acknowledgements** | Next Steps now is core-funded by the ESRC. |
| **Ethics** | Ethics approval is obtained for each follow-up from an NHS Research Ethics Committee (REC). In addition, separate REC approval is in place to cover the ongoing activities of the study in between major sweeps of data collection (i.e. keeping in touch with and tracing cohort members; cleaning, documenting and providing access to the data for research; and linking data from administrative sources to survey data to increase the utility of the data for research). |
| **Website for Data Requests** | <https://cls.ucl.ac.uk/cls-studies/next-steps/> |
| **COVID-19 survey dates** | Three surveys: May 2020, Sept-Oct 2020 and Feb-Mar 2021 |
| **Citation** | University College London, UCL Institute of Education, Centre for Longitudinal Studies. (2022). COVID-19 Survey in Five National Longitudinal Cohort Studies: Millennium Cohort Study, Next Steps, 1970 British Cohort Study and 1958 National Child Development Study, 2020-2021. [data collection]. 4th Edition. UK Data Service. SN: 8658, DOI: 10.5255/UKDA-SN-8658-4 |

| **NSHD: Medical Research Council National Survey of Health and Development** | |
| --- | --- |
| **Description of Study Population (including citations and references if required)** | The MRC National Survey of Health and Development (NSHD) is a socially stratified birth cohort of 2,547 women and 2,815 men. It is a sample of all births in England, Scotland, and Wales that occurred in one week in March 1946, and consists of all single births to married women with a husband in non-manual and agricultural employment and 1 in 4 of all comparable births to women with a husband in manual employment.^1^  ^1^Kuh et al. Cohort profile: updating the cohort profile for the MRC National Survey of Health and Development: a new clinic-based data collection for ageing research. Int J Epidemiol. 2011 Feb;40(1):e1-9. doi: 10.1093/ije/dyq231. |
| **Acknowledgements** | The UK Medical Research Council provides core funding for the MRC National Survey of Health and Development (MC_UU_00019/1). We are extremely grateful to the NSHD study members for their lifelong participation and continuing support; and to past and present members of the study teams, who helped to collect and process the data. |
| **Ethics** | Ethical approval for the study was obtained from the UK Research Ethics Committee (REC). |
| **Website for Data Requests** | <https://skylark.ucl.ac.uk/> |
| **COVID-19 survey dates** | Three surveys: May 2020, Sept-Oct 2020 and Feb-Mar 2021 |
| **Citation** | University College London, MRC Unit for Lifelong Health and Ageing. (2021). COVID-19 Survey in Five National Longitudinal Cohort Studies: MRC National Survey of Health and Development, 2020-2021: Special Licence Access. [data collection]. 3rd Edition. UK Data Service. SN: 8732, DOI: 10.5255/UKDA-SN-8732-3 |

| **Understanding Society – the UK Household Longitudinal Study** | |
| --- | --- |
| **Description of Study Population** (including citations and references if required) | Understanding Society, the UK Household Longitudinal Study, is a longitudinal survey of the members of ~40,000 households (at Wave 1, 2009-10) in the United Kingdom. The survey sample consists of a large General Population Sample (~26,000 households) plus three other components: the Ethnic Minority Boost Sample (~4,000 households), the former British Household Panel Survey sample (~8,000 households) and the Immigrant and Ethnic Minority Boost Sample (~2,900 households, added at Wave 6). Household and individual interviews are conducted annually. The study is multi-topic and multi-purpose.  From April 2020 to September 2021, participants from the main Understanding Society sample were asked to complete nine short web-surveys (with a telephone option in some months). The COVID-19 study covered the changing impact of the pandemic on the welfare of UK individuals, families and wider communities. ~18,000 individuals provided a full or partial interview at Wave 1 (April 2020).  At Wave 8 of the COVID-19 study, 8477 participants provided consent to link their survey data to administrative health records. |
| **Acknowledgements** | Understanding Society is an initiative funded by the Economic and Social Research Council and various Government Departments, with scientific leadership by the Institute for Social and Economic Research, University of Essex, and survey delivery by NatCen Social Research and Kantar Public  The COVID-19 study (2020-2021) was funded by the Economic and Social Research Council and the Health Foundation. Serology testing was funded by the COVID-19 Longitudinal Health and Wealth – National Core Study. Fieldwork for the web survey was carried out by Ipsos MORI and for the telephone survey by Kantar. |
| **Ethics** | The University of Essex Ethics Committee has approved all data collection on Understanding Society main study, COVID-19 surveys and innovation panel waves, including asking consent for all data linkages except to health records.  Approval for asking consent for health record linkage and for the collection of blood and subsequent serology testing in the March 2021 wave of the COVID-19 study was obtained from London – City & East Research Ethics Committee (21/HRA/0644). |
| **Website for Data Requests** | <https://ukllc.ac.uk> or <https://ukdataservice.ac.uk> |
| **COVID-19 survey dates** | Eight surveys: Apr 2020, May 2020, Jun 2020, Jul 2020, Sept 2020, Nov 2020, Jan 2021, Mar 2021 |
| **Citation** | University of Essex, Institute for Social and Economic Research. (2022). Understanding Society: Waves 1-12, 2009-2021 and Harmonised BHPS: Waves 1-18, 1991-2009. [data collection]. 17th Edition. UK Data Service. SN: 6614, <http://doi.org/10.5255/UKDA-SN-6614-18>.  University of Essex, Institute for Social and Economic Research. (2021). Understanding Society: COVID-19 Study, 2020-2021. [data collection]. 11th Edition. UK Data Service. SN: 8644, DOI: 10.5255/UKDA-SN-8644-11 |
